# Mapping patient journeys: a novel method to explore patient and carer experiences of injectable anticipatory medication care in the community and identify opportunities for improvement

**DOI:** 10.1101/2025.10.09.25336476

**Authors:** Rosanna Fennessy, Artemis Paterson, James Ward, John P. Clarkson, Ben Bowers

**Author notes:** corresponding author **Corresponding author information:** Address: East Forvie Building, Forvie Site, Addenbrookes Biomedical Campus, Robinson Way, Cambridge, CB2 0SZ.

## Abstract

**Background:** Injectable anticipatory medications are routinely prescribed ahead of need in many countries to help manage distressing end-of-life symptoms. However, little is known about the lived experience of patients and informal caregivers as they navigate the prescription and use of anticipatory medications.

**Aim:** To understand patient journeys in navigating anticipatory medication care, and to identify interactions with the greatest potential for improvement.

**Design:** Qualitative secondary analysis of longitudinal interview data using framework analysis and patient journey mapping techniques.

**Setting/participants:** Eleven patient-centred cases receiving end-of-life care in the community. Six patients, nine informal caregivers and five healthcare professionals took part (28 interviews).

**Results:** Patient journeys with anticipatory medications differed from intended pathways. Participants appreciated having access to injectable medications for future symptom control. However, there was suboptimal information exchange between patients, informal caregivers and healthcare professionals regarding their purpose and threshold for use. Navigating unfamiliar and complex end-of-life medication support pathways was more successful where patients could self-advocate or had live-in informal caregiver advocates, compared to those who lived alone or experienced communication difficulties.

**Conclusions:** Patient and informal caregiver experiences of timely symptom control could be improved by healthcare professionals having open and ongoing conversations about the role of anticipatory medications. Different patient contexts and fluctuating abilities point to a need for simplified and better signposted ways for accessing healthcare professional advice, and medication input. Journey mapping techniques offer a novel way to illustrate patient and informal caregivers lived experience and can be adapted for researching other pathways.

**Key Statements:** *What is already known about the topic?:*

- Anticipatory prescribing is considered best practice in aiding the timely control of distressing end-of-life symptoms in the community.
- The intended pathway for prescribing and using injectable anticipatory medications in the community is complex, consisting of many decision-making points and practical activities.
- Little is known about patients’ and families’ experiences of this care

*What this paper adds:*

- The use of journey mapping techniques illustrates how patients’ and informal caregivers’ experiences of navigating anticipatory medications differ from the intended pathway.
- Patient personas highlight how individual patient characteristics and advocacy skills greatly influence journeys with anticipatory medications.
- Patients and informal carers find the systems for using anticipatory medications inherently complex and clear professional signposting is needed.

*Implications for practice, theory or policy:*

- pen and ongoing conversations about the purpose and use of anticipatory medications need to be tailored to individual patient and informal carer contexts.
- experiential journey mapping techniques and personas is an innovative method for giving patients and their families a voice to help improve cross-organisational systems for delivering end-of-life care.

## Background

Injectable anticipatory medications are routinely prescribed ahead of need to help in managing any distressing end-of-life symptoms in the community.^1,2^ This is considered best practice in many countries, and as increasing numbers of people choose to die at home, their wider availability and use is promoted.^3–6^ Anticipatory medications are prescribed to alleviate symptoms of breathlessness, pain, nausea and vomiting, agitation and respiratory secretions,^7,8^ and to prevent unwanted emergency hospital admissions.^3,6,9^ They are typically prescribed by general practitioners (family physicians) and stored at home. The timing of prescription varies greatly, ranging from days to months ahead of likely need.^10–12^ This extended time frame is largely due to challenges of predicting end-of-life trajectories, particularly for people with non-cancer conditions such as multimorbidity and frailty.^13–15^ Decisions to subsequently administer medications are typically made by visiting district nurses and paramedics.^16–18^ Anticipatory medications are used in approximately 60% of cases where they are prescribed and are first started a median of 3 days before death.^16^

The intended injectable anticipatory medication pathway includes several key processes: prescribing, dispensing, storage, administration, and disposal after death; this involves multiple interactions with different healthcare services. Research has highlighted that patients and informal caregivers find having these injectable medications in the home both reassuring but also a cause of concern as they can be viewed as a harbinger of death.^6,17,19,20^ Informal caregivers also express concerns about the risks of medication causing over-sedation and hastening death.^6,19^ However, little is known about how patients and informal caregivers, in varying contexts, experience anticipatory medication care journeys.

Healthcare improvement initiatives are increasingly utilising systems approaches from engineering and inclusive design.^21–24^ These methods help to improve patient safety and performance by mapping and learning from patient-centred journeys and interactions with healthcare services, and by focusing on patients’ unique needs and capabilities when (re)designing healthcare systems.^24–26^ Tools include using ‘personas’ to represent different patient characteristics,^27–29^ alongside mapping ‘touch points’ and ‘pain points’ to identify important healthcare interactions and challenges within journeys.^22,24,28,30,31^ However, these methods and tools remain underused in healthcare research for illustrating and learning from lived experiences as people navigate intended care pathways.

Using secondary data from in-depth qualitative interviews, our study aims were to understand patient journeys in navigating anticipatory medication care, and to identify interactions with the greatest potential for improvement.

## Methods

### Design

Secondary qualitative analysis of longitudinal multi-perspective interview data, which was originally collected to explore views and experiences of anticipatory medication care. The study was guided by a constructivist approach, acknowledging that views and experiences are shaped through interactions and shared social worlds.^32^

### Ethical approval

The South Cambridgeshire Research Ethics Committee (reference: 19/EE/0361) (01/2020) granted approval for the original study, including the use of anonymised data for secondary analysis.

### Setting and participants

Data comprised eleven patient-centred case studies in two English counties. Patients were prescribed anticipatory medications prior to recruitment and lived at home or in residential care. Their informal caregivers and up to one healthcare professional involved in their care were recruited. In total, six patients, nine informal caregivers, three nurses and three general practitioners took part. All were aged over 18 years. Written informed consent was obtained during the primary study to allow anonymised data to be used for further research. One general practitioner did not consent to this, and their data has been excluded.

### Data collection

The dataset comprised experiential longitudinal data across multiple interviews collected May-December 2020. Interview guides explored participants’ perspectives on the prescribing of anticipatory medications and experience of use, in addition to preferences for future care (see Supplementary Document 1).

### Data analysis

Analysis comprised four sequential stages:

1. Interview transcripts were analysed using a framework analysis approach^33,34^ Analysis was undertaken by RF, a research psychologist, AP, a medical student, and BB, a clinical academic community nurse who conducted the original interviews. We inductively and deductively created codes related to patient context, and experiences of anticipatory medications and associated end-of-life care. We were also guided by concepts from patient safety mapping techniques including ‘touch points’ (interactions with healthcare professionals), perceived ‘pain points’ (bad experiences), and patient or carer ‘adaptations’. Using NVivo 14, RF and AP initially coded the same three cases independently and iteratively refined the coding framework through reflexive group discussions with the wider study team. Remaining transcripts were then coded, and the framework adapted as new codes were identified.
2. The eleven cases were systematically compared across codes including patient and informal caregiver capabilities, personal context, touchpoints and pain points, how far before death anticipatory medications were prescribed and their perceived usefulness. We selected three cases which highlighted the range of circumstances and experiences of those receiving anticipatory medications to create the personas Sue, Ted and Liam. Summaries of the remaining eight cases can be found in Supplementary Document 2. Our personas were derived from individual cases as we wanted to represent lived experience accurately and use verbatim quotations; however, each persona reflects characteristics of wider patient populations receiving end-of-life care in the community, including shorter or longer end-of-life trajectories, experiences of those living alone or with family, and variations in advocacy capabilities.
3. Together, RF, AP and BB visually mapped the three personas’ journeys, alongside the intended journey with anticipatory medications to illustrate similarities or differences (Supplementary Document 3). We identified three recurring factors which influenced these journeys and were reflected in all eleven patient cases.
4. The persona journeys were professionally illustrated to highlight these factors.

## Results

Injectable anticipatory medications were prescribed in all eleven patient-centred cases between 5 and 294 days before death or final interview. They were used in seven cases (64%) (Table 1).

**Table 1:**
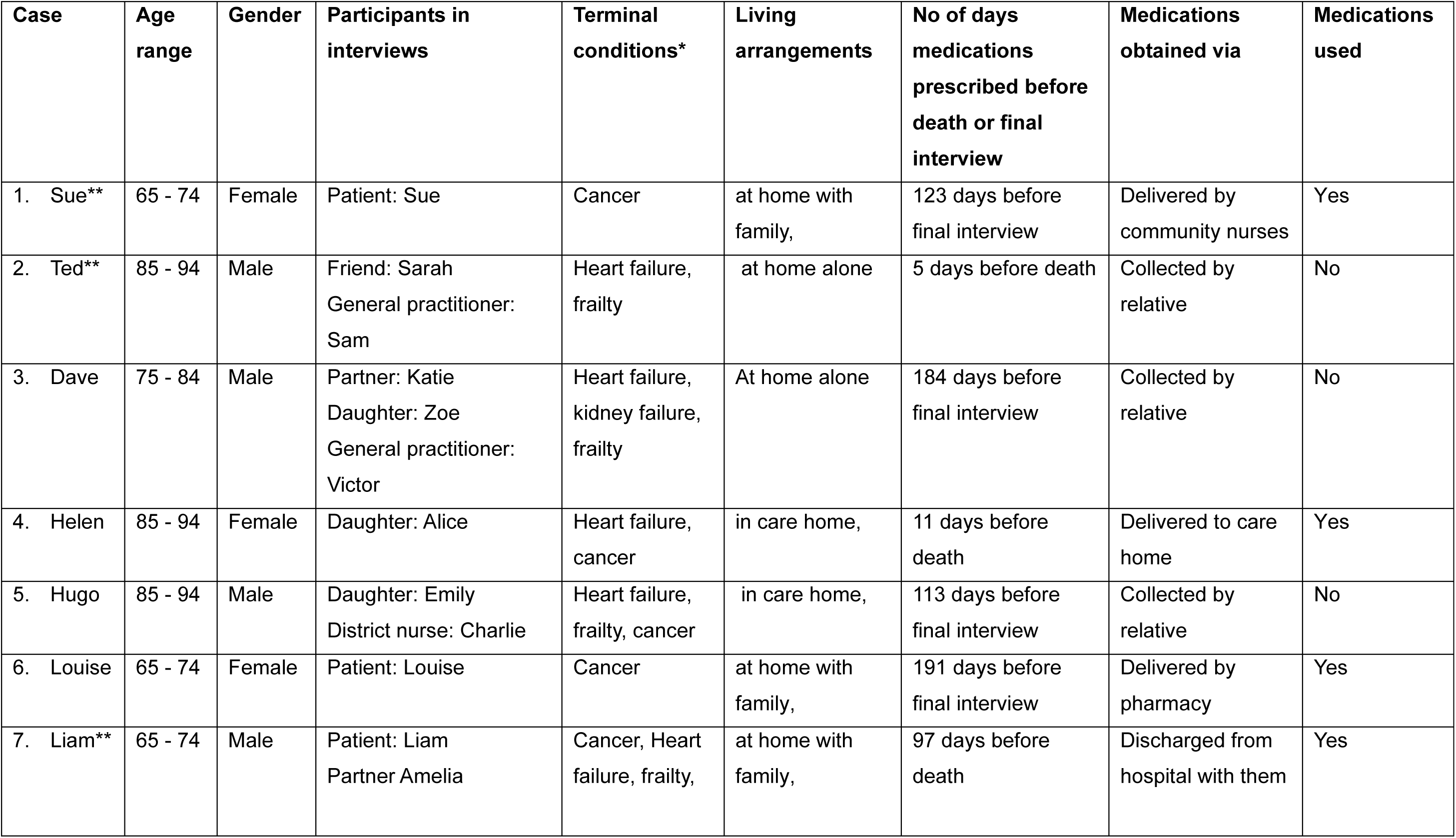
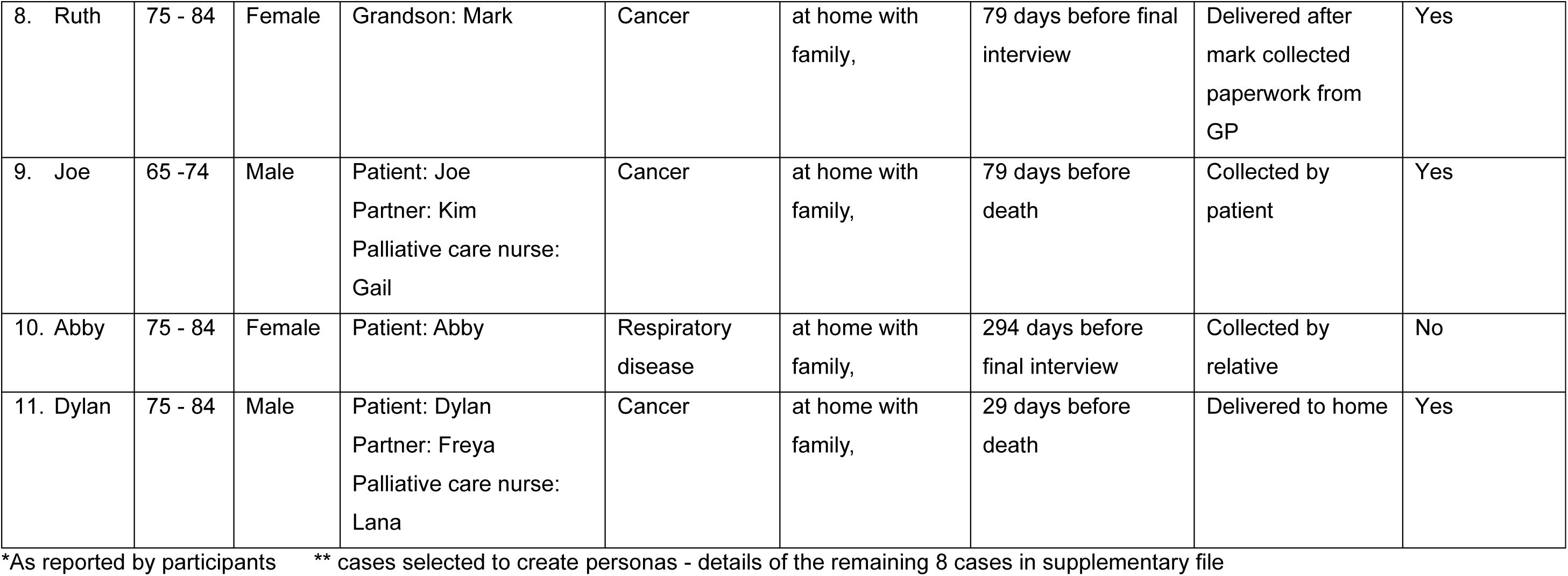
Participant details.

| Case | Age range | Gender | Participants in interviews | Terminal conditions* | Living arrangements | No of days medications prescribed before death or final interview | Medications obtained via | Medications used |
| --- | --- | --- | --- | --- | --- | --- | --- | --- |
| 1. Sue** | 65 - 74 | Female | Patient: Sue | Cancer | at home with family, | 123 days before final interview | Delivered by community nurses | Yes |
| 2. Ted** | 85 - 94 | Male | Friend: Sarah<br>General practitioner: Sam | Heart failure, frailty | at home alone | 5 days before death | Collected by relative | No |
| 3. Dave | 75 - 84 | Male | Partner: Katie<br>Daughter: Zoe<br>General practitioner: Victor | Heart failure, kidney failure, frailty | At home alone | 184 days before final interview | Collected by relative | No |
| 4. Helen | 85 - 94 | Female | Daughter: Alice | Heart failure, cancer | in care home, | 11 days before death | Delivered to care home | Yes |
| 5. Hugo | 85 - 94 | Male | Daughter: Emily<br>District nurse: Charlie | Heart failure, frailty, cancer | in care home, | 113 days before final interview | Collected by relative | No |
| 6. Louise | 65 - 74 | Female | Patient: Louise | Cancer | at home with family, | 191 days before final interview | Delivered by pharmacy | Yes |
| 7. Liam** | 65 - 74 | Male | Patient: Liam<br>Partner Amelia | Cancer, Heart failure, frailty, | at home with family, | 97 days before death | Discharged from hospital with them | Yes |
| 8. Ruth | 75 - 84 | Female | Grandson: Mark | Cancer | at home with family, | 79 days before final interview | Delivered after mark collected paperwork from GP | Yes |
| 9. Joe | 65 -74 | Male | Patient: Joe<br>Partner: Kim<br>Palliative care nurse: Gail | Cancer | at home with family, | 79 days before death | Collected by patient | Yes |
| 10. Abby | 75 - 84 | Female | Patient: Abby | Respiratory disease | at home with family, | 294 days before final interview | Collected by relative | No |
| 11. Dylan | 75 - 84 | Male | Patient: Dylan<br>Partner: Freya<br>Palliative care nurse: Lana | Cancer | at home with family, | 29 days before death | Delivered to home | Yes |

\*As reported by participants
\*\* cases selected to create personas - details of the remaining 8 cases in supplementary file

Recurring factors impacting patient journeys:

1. *Perceptions of anticipatory medications* Patients and informal caregivers noted the usefulness of anticipatory medications to relieve potential pain and suffering in the future. However, when medications were used, they were not always perceived as being helpful. Some patients and informal caregivers had concerns about medications causing over-sedation and potentially hastening death, others found that hospitalisation was more helpful in managing symptoms.
2. *Quality of information exchange* Information sharing between healthcare professionals and patients and informal caregivers shaped patient journeys, both positively and negatively. This included communication about prescribing anticipatory medications and when they might be used, along with ongoing contact and dialogue as patients’ journeys progressed.
3. *Ability to navigate healthcare systems* Navigating complex and unfamiliar processes related to anticipatory prescribing and end-of-life care was often problematic. Patients who could advocate for themselves or had live-in caregivers were more successful in accessing timely care, compared to those who lived alone or had less capacity to communicate.

These factors are illustrated in the journeys of our three patient personas.

### Sue’s journey

**Persona: Sue, 74 years old**

**Terminal condition: cancer**

**Anticipatory medications prescribed 123 days before the final research interview**

Sue, a former nurse, had been living with cancer for several years. She lived with her husband Marcus. Her health was deteriorating slowly, including becoming almost blind and deaf. However, she remained fiercely independent, managing her daily life and healthcare interactions herself.

Sue wanted to stay at home for as long as possible but did not want her husband to become her carer.

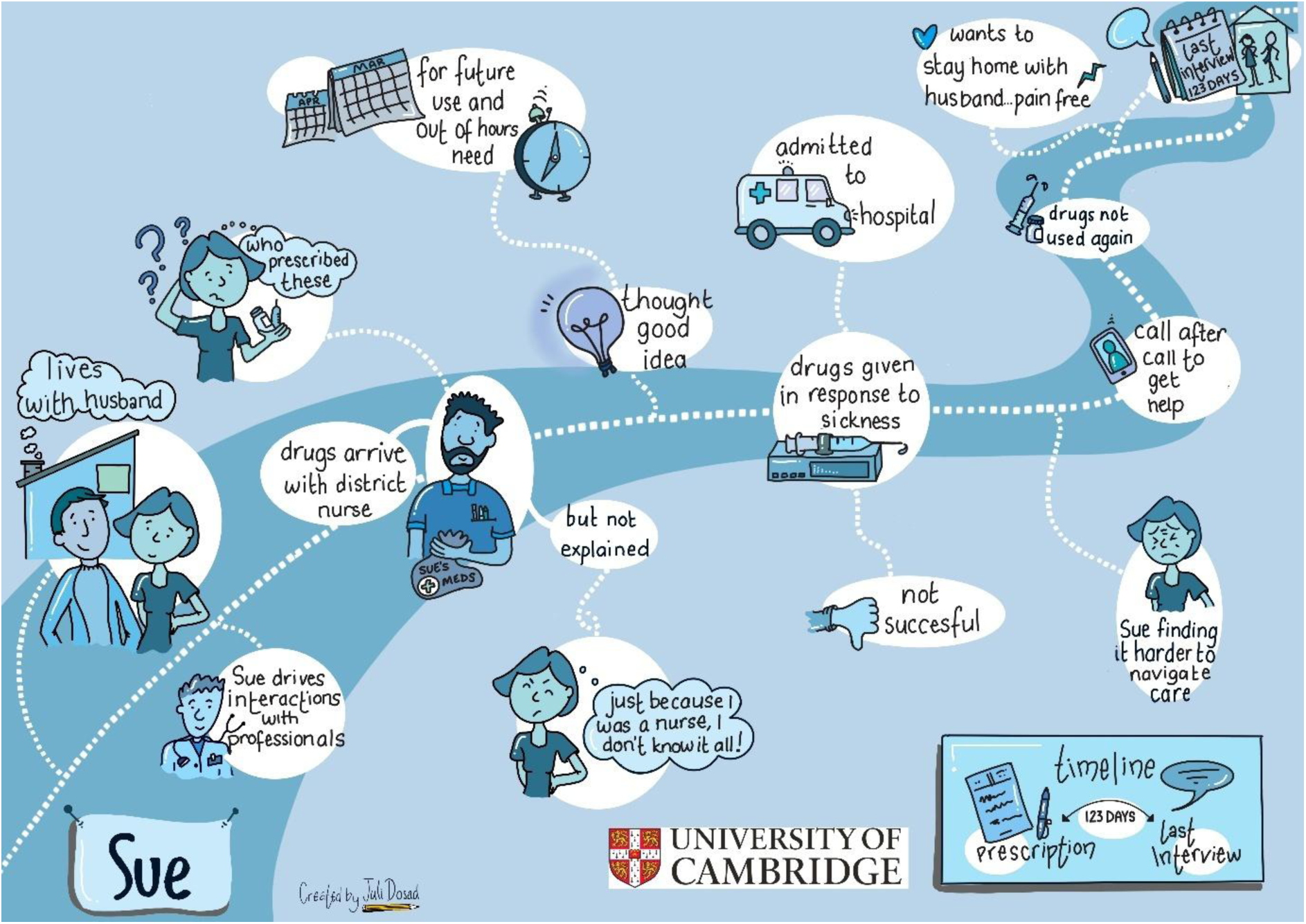

When Sue’s oncologist advised that care was now palliative, anticipatory medications were prescribed, although she was not sure by whom - they “just appeared” [Sue] with the district nurses. Sue read the leaflets in the packets but thought that the lack of explanation from healthcare professionals was unhelpful and might be due to assumptions about her medical background:

> *“I don’t want people to think that I know things because I don’t… I do think sometimes they presume that they don’t have to say things because you’re a nurse and you should know that but it don’t work like that!” [Sue]*

Nevertheless, Sue was happy to have anticipatory medications in the house for potential future use:

> *“I know what they’re all for but I think, you know, you’ve just got to have them quick, if you need them, especially at weekends and stuff like that.” [Sue]*

Sue advocated for herself with healthcare professionals, telling the district nurses that she would contact them if necessary. However, she expressed concerns that her general practitioner was unfamiliar with her situation and needs:

> *“She [the general practitioner] didn’t know anything about me and then I just said to her about this and she said, oh you’ve had breast cancer in the past. I said, oh that was ages ago, I gave her a run down and everything but she didn’t really know anything about me.” [Sue]*

Sue experienced an episode of continuing sickness, and the anticipatory medications were commenced using a syringe driver. However, after a district nurse and two out-of-hours doctors attended, she was admitted to hospital. Sue subsequently felt that the anticipatory medications were not helpful:

> *“I’d probably say, well, it just didn’t go right for me. Or perhaps I was just too far gone for anything to, I needed fluid… It was put up in the ambulance, I was so dehydrated… Having the drugs, without having fluid first, I think it was just a bit of a mess up.” [Sue]*

On returning home, Sue found navigating health care services increasingly challenging. She had difficulty with medication-related constipation:

> *“The bad days, just too much happening and too much messing about with phone calls, trying to get somewhere. Like Saturday morning, because I was a bit desperate with this bowel thing, I was just like, it was one thing after another, 111, through to the chemists, chemists computers are down. And then when you’re doing all this, you’re finding that, oh my goodness, I haven’t eaten lunch and I’ve got to take pills and it’s getting late.” [Sue]*

Sue continued to deteriorate and was almost blind, however she wanted to remain independent. At the end of the second interview, she was still at home and the anticipatory medications had not been used again. When asked about her wishes for the future she commented:

> *“All I want to do is just go to sleep and not wake up. So I’m just comfortable and, touch wood, as I say, I’m not in any pain, but I just want to be comfortable and go to sleep, and that’s it.” [Sue]*

### Ted’s journey

**Persona: Ted, 91 years old**

**Terminal conditions: heart failure and frailty**

**Anticipatory medications prescribed 5 days before Ted’s death**

Ted lived alone and was previously independent, driving and cooking for himself. He liked a beer and enjoyed visits from his good friend Sarah (a retired nurse) and her dog.

He rapidly declined in health and became bedbound after returning home from respite care in a nursing home. He struggled to take oral medication due to swallowing difficulties. Sarah and Ted’s niece took turns to visit daily. Professional carers attended each day to provide personal care. District nurses visited periodically to review his needs.

Ted wanted to be comfortable and without pain in his last days of life.

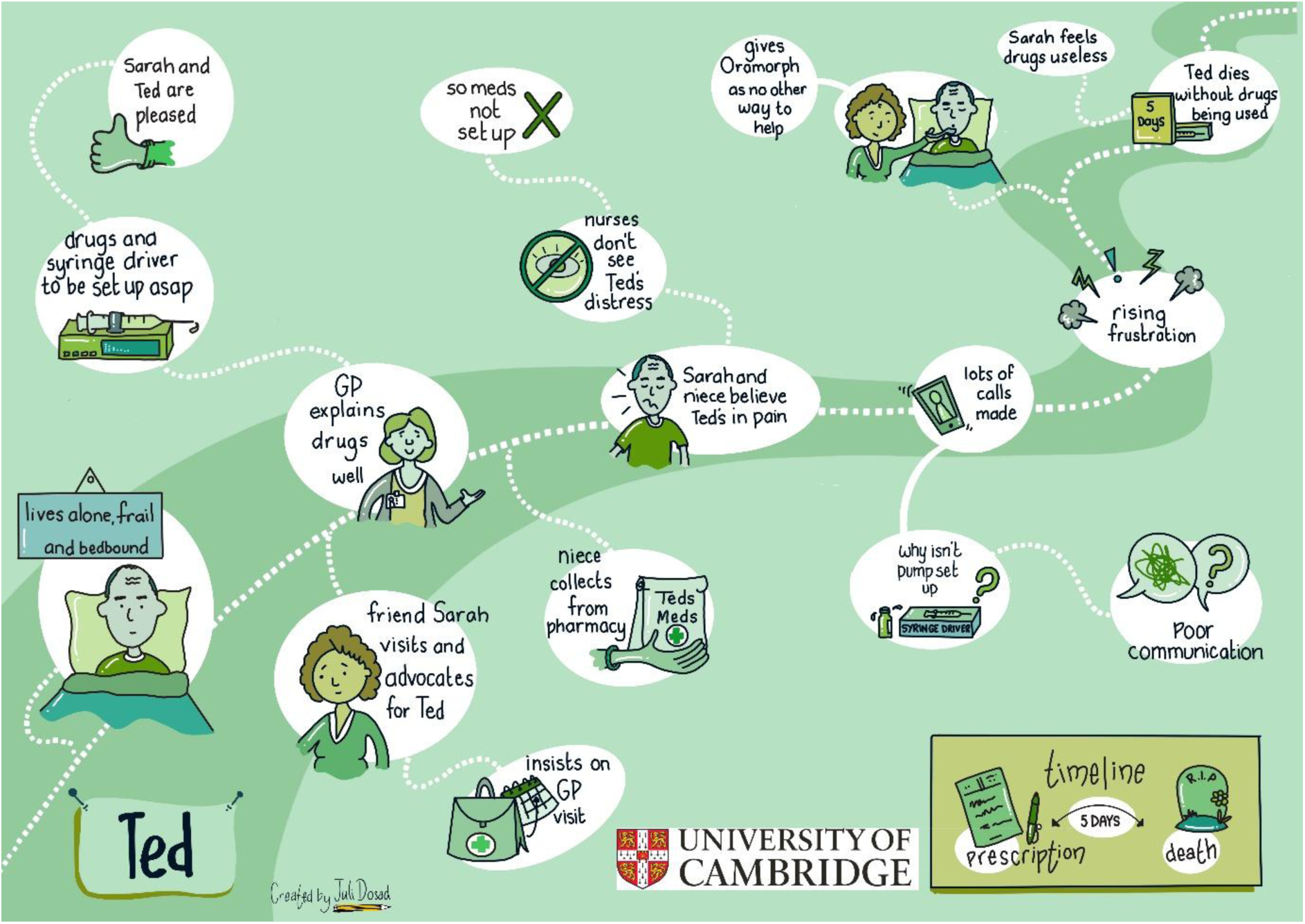

Sarah’s nursing background and intuition led her to insist on a general practitioner home visit for Ted to discuss his deterioration and the likelihood that he was now dying. Sarah and Ted’s niece were present at this visit and advocated on Ted’s behalf:

> *“I just felt he’s just been left and if I hadn’t had that knowledge and knew what I wanted for him or what was available for him, I don’t think he would have had it.” [Sarah]*

The general practitioner explained what the anticipatory medications were for and prescribed them to start via a syringe pump. They believed this would be used immediately to help with Ted’s pain:

> *“And she [the general practitioner] said, well we’re going to give you some things to make you comfortable but it’s not going to make it any quicker but it’ll make that time, hopefully, more comfortable for you… She implied that we would be starting it there and then because Ted wants to die, he’s uncomfortable, he was being sick and he was happy to start that pump there and then.” [Sarah]*

Sarah thought the syringe pump would be particularly useful for managing Ted’s nausea and chest secretions:

> *“Well to me the pump’s just going to make him more comfortable, and he keeps trying to be sick, we try and give him things and he’s had a lot of secretion that he keeps coughing up, you know what I mean, and it will hopefully just let him rest and have nice dreams and not be uncomfortable.” [Sarah]*

The general practitioner identified a pharmacy with injectable medication stock and Ted’s niece collected them. However, they weren’t given any further details of the drugs or next steps. The syringe pump was not set up and no medication was given by the nurses:

> *“I think there should have been some sort of conversation with us about them, we weren’t given a leaflet about it, or how it would all be set up, we weren’t informed of anything like that. So because we were led to believe by the GP that the palliative nurses will come up and set this pump up and it’ll be nice and relaxed and that, that’s what we were thinking would happen which it never did.” [Sarah]*

Sarah and Ted’s niece found getting information from Ted’s healthcare team challenging and were distressed that they could not persuade the district nurses to start the anticipatory medications. Their daily visits did not coincide with the nurses, limiting opportunities to communicate:

> *“We’ve phoned the palliative nurses but what you do, you go through a call centre or whatever. We’ve said look, he’s at home on his own, these palliative nurses are coming in, we don’t know what they’re doing… There’s nothing written at home to say what the plan is, so we feel we’re not in the loop… Why has the pump not been done and it is just very frustrating, you know, to see him like that and stuff’s there to make him more comfortable and nothing’s happening.” [Sarah]*

The nurses told Sarah by phone that Ted did not need the anticipatory medications. However, Sarah observed Ted in distress on her morning visits and gave him liquid morphine to make him more comfortable for when the carers arrived to wash him. She left notes for the nurses to let them know this and rationalised that maybe they did not witness his distress:

> *“Maybe the district nurses or palliative care nurses would come in, and he’d seem fine to them, because they only saw him for like a five-minute slot. But they didn’t see how he was all distressed and hallucinating and things like that, which was, it was worse for me because I was the one to find him.” [Sarah]*

An interview with Ted’s general practitioner highlighted differences in healthcare professionals’ perceptions over whether the injectable medications should have been started:

> *“I prescribed it so it could be delivered by syringe driver as well as for if needed. And I sort of, I thought that was going to be the way to go… but we refer to the district nurses… they did go and visit him twice a day but they didn’t think a driver was necessary. So that was a slight discrepancy in what I thought would happen and versus what did happen.” [Sam, general practitioner]*

Ted died at home five days after the medications were prescribed, without any of the drugs being given. Sarah considered that the anticipatory medications were “useless because nobody would give him anything.” [Sarah].

### Liam’s journey

**Persona: Liam, 73 years old**

**Terminal conditions: cancer, heart failure**

**Anticipatory medications prescribed 97 days before Liam’s death.**

Liam lived at home with Amelia, his partner and carer. His health was declining slowly, and he had been in and out of hospital many times with various infections while receiving palliative chemotherapy. Amelia had taken over overseeing Liam’s oral medication and giving his insulin to help manage his diabetes.

Liam wanted to stay at home and avoid more hospital stays. He didn’t want to be in pain but did not want to be medicated like a ‘zombie’.

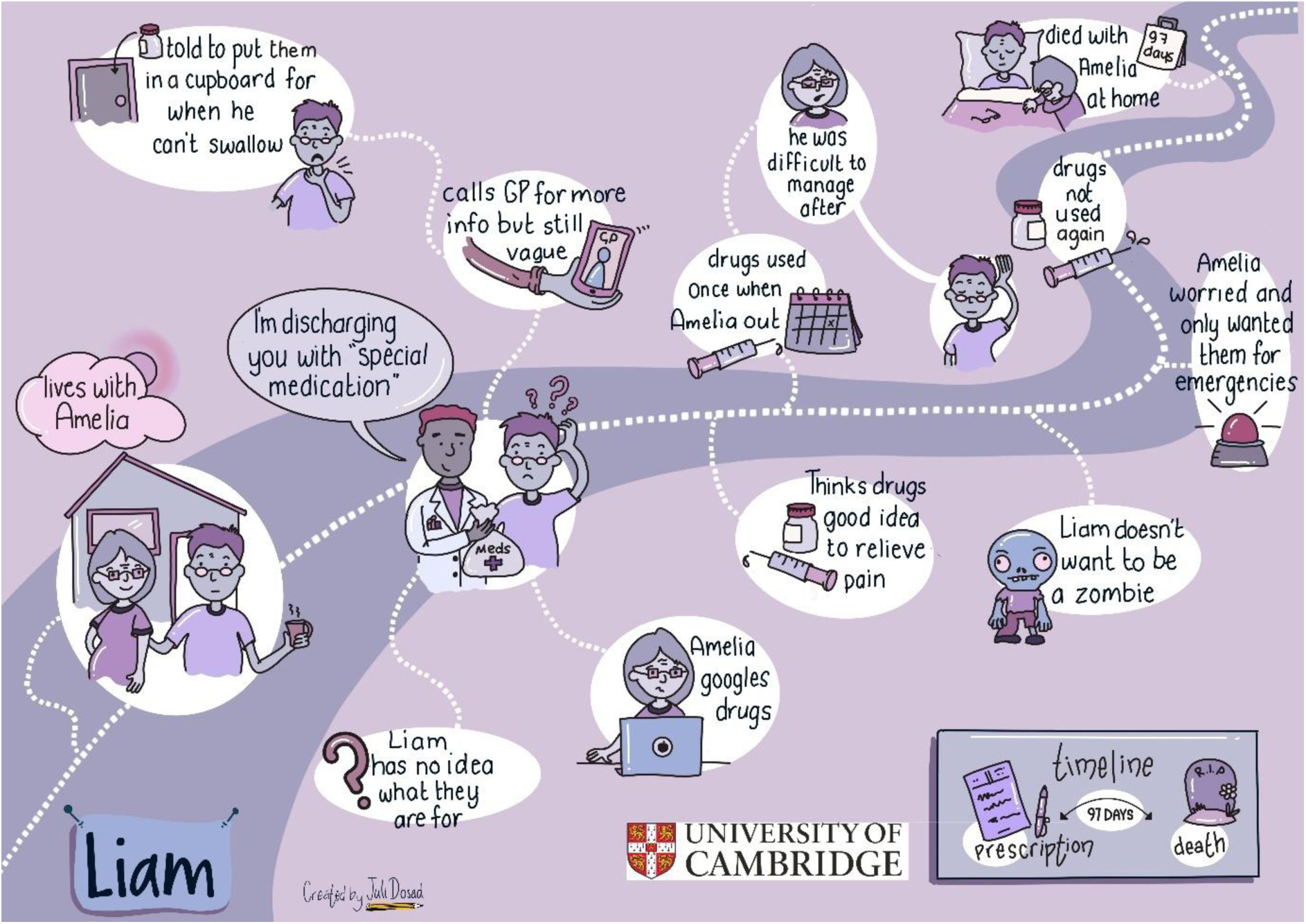

At the end of a period in hospital, Liam’s discharge was delayed as some “special medications” [Liam] had been prescribed but not dispensed. Liam asked questions about what turned out to be the anticipatory medications, but the hospital pharmacist did not explain them:

> *“I said, “Well, what is this?”, and they said, “We don’t know, it’s some special medication that the doctors have to send, you have to go home with”, which apparently, learning, on reflection, is this bag of goodies [anticipatory medication]. So that was the first thing I knew about it. No one had mentioned it to me, I’d never heard of it.” [Liam]*

Once home, Amelia looked up the names of the medications on the internet. She also phoned their general practitioner, although she would have preferred more detail in his response:

> *“I said to Dr Potter, “Well, I’ve got a bag here and it says Just in Case”, and he said, “Yes, put them in the cupboard for later in case Liam can’t swallow”…, so he sort of like skirted over but told me what they’re for.” [Amelia]*

Amelia advocated strongly on Liam’s behalf including his desire not to go back into hospital. They also believed the chemotherapy tablets were causing much of Liam’s pain, and managing this became a source of concern:

> *“I’ve got… cancer. Which no ones used the word terminal to me, but I know it’s terminal, okay. One or two doctors have said, “Yeah, we can’t, we can’t cure it, Liam, but we just, we can control it”… If controlling it means suffering pain all the time, then I’m not interested in that. But I don’t want to go to the other side so that I’m sort of no pain but just lying like a zombie in bed, I can’t see any point in having that sort of life. So we’ve got to find a balance between the two, and it’s just finding that balance seems to be the struggle at the moment.” [Liam]*

About seven weeks before his death and whilst Amelia was out, a district nurse gave Liam a dose of the anticipatory medications to “calm him” [Amelia]. Amelia was upset by its impact as she was unable to manage Liam physically afterwards and was concerned about the potential for the drugs to be misused. She subsequently insisted they only be used in emergencies. Liam died at home comfortably with Amelia by his side. The anticipatory medications were not used again:

> *“It was reassuring that if he did suffer, then I knew that all I had to do was to call a district nurse and she would come out and take that suffering away from him, that was always there in my mind…. When I had the episode… I thought how easy it is for someone just to come and, in my terms, bump somebody off. So, yeah, no, they stayed in the cupboard, for purely an emergency only.” [Amelia]*

### Identifying interactions with the greatest potential for improvement

We identified three recurring opportunities where interactions between patients, informal caregivers and healthcare professionals offered potential for improvement.

*1. Explanations of anticipatory medications*

Anticipatory prescriptions need open and tailored explanations by healthcare professionals. This includes exploring patients’ and informal caregivers’ interpretations of the purpose of medications. The three persona journeys highlight that anticipatory medications were often not adequately explained. For example, Amelia had to source information from the internet. Those with medical backgrounds had some understanding, but as noted by Sue, she did not want people to make assumptions about her knowledge. While participants commented on the usefulness of these medications for the relief of future pain, further information and discussion were often needed, as differing perceptions about the medications and their effects were commonplace.

2. *Optimising system transparency*

Although the decision to prescribe anticipatory medications was driven by healthcare professionals, greater transparency and ongoing communication could have helped patients and informal caregivers to better navigate support systems related to their use. This was particularly notable for Ted and Sarah who didn’t understand why the medications and syringe pump were not used. The challenges of communicating with multiple healthcare teams also impacted on experiences of using anticipatory medications. Sarah thought the medications were “useless’” due to the difference between the perceptions of the general practitioner and the nurses regarding the threshold for their use. For Amelia, anxiety regarding their potential to hasten death meant she no longer wanted them to be used. Continued dialogue and clear signposting regarding how to access help with symptom control could have helped alleviate patient and informal caregiver distress.

3. *Tailoring input to individual patients’ and informal caregivers’ circumstances*

Our three journeys illustrate how the different circumstances and resources of patients and informal caregivers influenced their ability to navigate unfamiliar healthcare systems. Sue maintained her self-advocacy, although found things more challenging over time, while Liam and Ted relied on others. However, Amelia was more successful in advocating, perhaps because she lived with Liam. Conversely, Sarah had considerable difficulty advocating for Ted’s ongoing needs. Recognising differences in the individual resources of patients and their informal caregivers over time, along with adapting care provision to meet variable situations and needs, would improve experiences of timely symptom control care.

## Discussion

Our study provides important insights into lived experiences of the prescription, dispensing and use of anticipatory medications. Participant journeys highlighted how patient and informal caregiver experiences varied from intended pathways. There was considerable variation in how far in advance of anticipated and actual need prescriptions were put in place, mirroring the findings of previous studies.^9,10,16^ Differences in patients’, informal caregivers’ and healthcare professionals’ perceptions regarding the purpose of anticipatory medication, and when they would be used, was a source of concern and frustration. Patients’ self-advocacy skills, alongside whether they lived with their informal caregivers, also influenced their ability to access and navigate unfamiliar and complex end-of-life care support.

### What this study adds

Having anticipatory medications available in the home is perceived to be reassuring for all involved,^2,19,35,36^ and our findings generally concur with this. However, we found that perceived gaps in information sharing regarding the purpose, safety and likelihood of use of these medications, caused concern to patients and informal caregivers, and may add to the psychological distress commonly experienced by patients and families at the end of life.^3,37^ There is a need for open and ongoing conversations about the role and purpose of anticipatory medications to explore and address any concerns.^12,19,38^ These need to be initiated by healthcare professionals and tailored to patients and informal caregivers existing knowledge and preferences for information.

Ongoing information sharing about symptom control is important as patients and informal caregivers may not recall conversations or retain information in stressful and ever-changing situations.^19,38–40^ Information sharing at the end of life is shaped by many factors including healthcare professional concerns about causing potential distress and their confidence and experience in having these conversations.^41–44^ Not all patients and families wish to engage in explicit conversations about what the future may hold.^39,45^ However, previous research suggests patients often have a preference for open and honest communication, ^39,46–48^ which reflects the desire for information found in our current study. Further research exploring generational and cultural preferences for information sharing is required.

Our patient journeys reflect differences in patient resources and self-advocacy skills; and informal caregivers became increasingly important in helping navigate anticipatory medication pathways. The work of informal caregivers is essential in meeting patients’ end-of-life care needs in the community, especially for those who live alone. They also assume significant responsibilities for managing medication and providing timely symptom control. ^35,38,49–52^ However, we found that informal caregivers did not feel sufficiently involved in decisions to use anticipatory medications, despite this being found to be empowering for all concerned.^49,53^ In our current study, this could have been facilitated by providing simplified methods to contact healthcare professionals for timely clinical medication advice which would have improved participants’ experiences.

Mapping patient journeys has become an important method to improve patient safety and healthcare services.^21–23^ However, the focus is often on mapping processes rather than patient experience and priorities. By adopting ideas from inclusive design techniques, such as creating personas, our novel use of secondary data enabled us to visually map patient journeys against intended pathways. This process exposed several ‘pain points’ for participants as they navigated the prescription and use of anticipatory medications, highlighting the need for greater flexibility and responsiveness in existing systems. Although this method of mapping experience needs further development, such as ensuring personas reflect a full range of diverse characteristics, it offers rich patient-centred insights which in future research may stimulate cross-organisational improvements in care.

### Strengths and limitations

Our qualitative analysis incorporated framework analysis and journey mapping techniques which allowed for rich description and visual illustration of patient and informal caregiver experiences. Our analysis was grounded in reflexivity, and we discussed our early findings with our Public and Clinician Advisory Group to incorporate their views on the implications for practice. However, as this was a secondary analysis, we were unable to ask participants follow up questions.

The patient personas we created represented the range of characteristics and circumstances from our sample, but these do not fully reflect the diversity of individuals who navigate end-of-life care services. We are currently seeking out unheard voices to ensure under-served populations are better represented in a new qualitative study. Similarly, while our secondary analysis incorporated data collected from healthcare professionals involved in prescribing the anticipatory medications, we were unable to triangulate our analysis of patient and informal caregiver experiences with the experiences of nurses involved. The original data was collected during the Covid pandemic so there may have been unique circumstances affecting patient journeys at this time. However, studies undertaken before the pandemic have highlighted similar difficulties with information exchange, shared decision-making and navigating end-of-life care pathways.^2,35,40^

## Conclusions

Injectable anticipatory medications are perceived to help in providing timely symptom control for patients dying in the community. Our novel study demonstrated that patient and informal caregiver experience of their prescription and use often differed from the intended pathway. Describing patient journeys with personas illustrated the challenges experienced in navigating unfamiliar end-of-life care systems. They highlight the need for open and ongoing dialogue initiated by healthcare professionals regarding anticipatory medications, to reduce patients’ and informal caregivers’ concerns and facilitate access to timely symptom control.

## Data Availability

The anonymised quantitative data used in this study may be requested by researchers through contacting RF or BB, first and last authors. Due to the sensitive subject nature of the anonymised qualitative data, they may be made available to researchers with evidence of a study protocol and Research Ethics Committee approval, and on completion of a data use and sharing agreement

## Author contributions

RF, AP, BB, JW and JC designed the study. RF, AP and BB conducted the data analysis. BB collected the original data. All authors contributed to the interpretation of the results. RF drafted the manuscript. All authors reviewed and revised the manuscript and approved the final version for submission. RF is the guarantor. The corresponding author attests that all listed authors meet authorship criteria and that no others meeting the criteria have been omitted. The lead author (RF) affirms that the manuscript is an honest, accurate and transparent account of the study being reported.

## Research ethical approvals

The study was approved by the South Cambridgeshire Research Ethics Committee (reference number: 19/EE/0361) 01/2020

## Acknowledgements

We would like to thank all the participants who gave up their valuable time and shared their experiences and insights; the members of our Public and Clinical Advisory Group for their invaluable input in helping to interpret the relevance of the findings; Juli Dosad for her professional illustrations of our patient journeys; James Brimicombe, Senior Data Manager for the Primary Care Unit, Department of Public Health and Primary Care, University of Cambridge, for his help with secure data storage; Angela Harper for her considerable administrative support; the National Institute for Health and Care Research Clinical Research Network (NIHR CRN) for their study support; Hertfordshire Community NHS Trust, Cambridgeshire and Peterborough NHS Foundation Trust and the other research sites for facilitating the research.

