## Supplemental file 1 for "Mapping patient journeys: a novel method to explore patient and carer experiences of injectable anticipatory medication care in the community and identify opportunities for improvement"

### Simultaneously reassuring and unsettling: a longitudinal qualitative study of community anticipatory medication prescribing for older patients

#### Supplementary file 1.

Initial interview schedule guides. Questions were continually reshaped in response to accounts, including previous interviews.

#### Patient and Informal Caregiver Participant Interview Schedule One

**JUST IN CASE DRUGS STUDY**

INTERVIEW SCHEDULE: PATIENT AND / OR INFORMAL CAREGIVER (FAMILY OR FRIEND) INTERVIEW ONE

Note: 1. Questions will be continually reshaped in response to data from previous interviews.

2. If patients or informal caregivers are interviewed individually, the questions will be phased in a suitable way to explore their views and experiences.

**Introduction:**

- Introduce yourself.
- Discuss the purpose of the study (the purpose of this research is to find out about patients and their family / friends opinions and experiences of 'Just in Case drugs' over time. We are also interested in **understanding patients and informal caregivers** involvement in the discussion and decision to prescribe and administer Just in Case drugs.
- Clarify that there are no right or wrong answers.
- We can pause or stop the conversation at any time, please just say if you would like to.
- Please ask for explanations of any questions whenever you want.
- Our conversation will be recorded with your permission.

**Explain that there are 4 themes the conversation is going to cover:**

- We will discuss the Just in Case drugs you / (patient's name) have been prescribed
- How you felt about these drugs being prescribed
- If you felt that you were given enough information about the drugs
- What you consider to be important in your / (patient's name) future care

**Semi-structured interview questions**

***Prompt: turn recorders on***

1. **Can you start by telling me about how your / (patient's name) health has been in the last three months?**

*Prompts:*

- *Have you / has (patients name) been having any treatment?*
- *Do you / does (patient's name) have problems with any symptoms?*
- *Are any healthcare professionals involved in your / (patient's name) care?*

2. **Could you tell me about the medications that you / (patient's name) are currently prescribed?**

   Ask what they call the drugs?

*Prompts:*

- ***If Just in Case drugs are not included in the list probe for any other drugs that have been prescribed/ are in the house but not currently being used?***
- *When were the Just in Case drugs prescribed and by whom?*
- *What are the drugs and what are they are for?*
- *Under what circumstances do you anticipate that they will be used?*
- *Where are they kept in the home?*

3. **How do you feel about having the Just in Case drugs in the home?**

*Prompts:*

- *Were you involved in getting the drugs from the chemists?*
- *If so, was it easy to get the drugs?*
- *Did you have any concerns about having them in the home?*

- *Were you reassured to have them in the home?*

4. **How involved did you feel in the discussion about prescribing these drugs?**

*Prompts:*

- *Did anyone explain what they might be used for?*
- *Was the information and the way it was supplied helpful?*
- *Was there information that was not helpful?*
- *Was there information not talked about that you would have found helpful to know?*

5. **Did the healthcare professional who prescribed the Just in Case drugs ask you if you wanted them prescribing?**

*Prompts:*

- *If gives a yes or no answer: please could you explain a bit more*
- *How you felt about them being prescribed?*

**Thank you, that helpful to know.**

I'd like to explore (patients name) care to understand what's important to you.

6. **Thinking ahead, what do you consider important in your / (patient's name) future care?**

*Prompts:*

- *Follow where the participant/s wants to go, exploring their care wishes and understanding of their situation.*
- What is important for you in supporting your / (patient's name) comfort?
- If the participant/s raises that drugs are for symptom control – are there symptoms they are concerned may develop?
- Does the participant / do the participants expect Just in Case drugs to be used at some point?

7. **I think that's everything I wanted to talk about, do you have anything else you would like to say or pressing thoughts that I have not asked you?**

**Close the interview**

#### Patient and Informal Caregiver Participant Interview Schedule Two

**JUST IN CASE DRUGS STUDY**

INTERVIEW SCHEDULE: PATIENT AND / OR INFORMAL CAREGIVER (FAMILY OR FRIEND) INTERVIEW TWO

Note: 1. Questions will be continually reshaped in response to data from previous interviews.
2. If patients or informal caregivers are interviewed individually, the questions will be phased in a suitable way to explore their views and experiences.
3. The second interview will build on and refer to what was established in the first one.

**Introduction:**

- The purpose of this second research conversation is to find out about your opinions and experiences of 'Just in Case drugs' now you have had some / (patients name) has some administered.
- Clarify that there are no right or wrong answers.
- We can pause or stop at any time, please just say if you would like to.
- Please ask for explanations of any questions whenever you want.
- Our conversation will be recorded with your permission.

**Explain that there are 3 broad themes the conversation is going to cover:**

- What has changed for you / (patient's name) since our first conversation
- Your experiences of the Just in Case drugs being used
- What you consider is important in supporting your / (patients name) comfort now

**Semi-structured interview questions**

***Prompt: turn recorders on***

1. **Can you start by telling me what has changed for you / (patient's name) since we last met?**

*Prompts:*

- *Have you / has (patient's name) had problems with any symptoms in the last few days?*
- *Which healthcare professionals are now involved in your / (patient's name) care?*

2. **You have / (patient's name) has had some of Just in Case drugs given by a nurse or doctor. Would you mind telling me about this experience?**

*Prompts:*

- *How did the drugs come to be used?*
- *How did you find the process of getting someone to come and give the drugs?*
- *If they were called for help, how quick were professionals in responding / coming?*
- *Who made the decision to give the drugs?*
- *Did you feel you were involved in the decision to give the drugs?*

3. **Was any information about the drugs being administered discussed with you?**

*Prompts:*

- *Did a nurse or doctor explain what they were for and their effects before they administered them?*
- *Did anyone explain about the drugs and their effects after they were administered?*
- *Was the information and the way it was supplied helpful?*
- *Was there information that was not helpful?*
- *Was there information not talked about that you would have found helpful to know?*
- *If a syringe driver was set up, did anyone discuss what it was for with you?*

4. **Did the drugs help or hinder you / (patient's name) in any way?**

*Prompts:*

- *H*ow effective do you feel the drugs were?

5. **How do you feel about these drugs being used?**

*Prompts:*

- *If a syringe driver was set up, how did you feel about this?*
- *Does the participant / do the participants expect Just in Case drugs to be used again?*
- *How do they feel about that?*

6. **Has having the Just in Case drugs affected how you see the future, or what is likely to happen in future?**

*Prompts:*

- *If yes, how has it affected how you see the future?*
- *Are you happy to have drugs in the house 'Just in Case' or does this cause you anxiety?*

**Thank you for sharing your thoughts and experiences.** I'd like to explore your / (patients name) care to understand what's important to you.

7. **What is important in supporting your / *(patient's name)* comfort now?**

*Prompts:*

- *Follow the participant/s prompts, exploring their care wishes and understanding of their situation.*
- *If the participant/s raise that (patient's name) is becoming much more unwell, give them the opportunity to talk about what they expect to happen, their hopes and concerns.*
- *Do you have any hopes or concerns about your / (patient's name) comfort in the coming days or weeks?*
- *Do you feel Just in Case drugs will be helpful in making it possible for you / (patient's name) to stay at home? (if the participant/s raise this as something that is important to them)*

8. **I think that's everything I wanted to talk about, do you have anything else you would like to say or pressing thoughts that I have not asked you?**

**Close the interview**

#### Patient and Informal Caregiver Participant Interview Schedule Three

**JUST IN CASE DRUGS STUDY**

**INTERVIEW SCHEDULE: PATIENT AND / OR INFORMAL CAREGIVER INTERVIEW THREE**

Note: 1. Questions will be continually reshaped in response to data from previous interviews.
2. If patients or informal caregivers are interviewed individually, the questions will be phased in a suitable way to explore their views and experiences.
3. This follow-up research interview will build on and refer to what was established in the previous ones.

**Introduction:**

- The purpose of this follow-up research conversation is to find out about your opinions and experiences of 'Just in Case drugs' and if they have changed since our previous meeting/s.
- Clarify that there are no right or wrong answers.
- We can pause or stop at any time, please just say if you would like to.
- Please ask for explanations of any questions whenever you want.
- The conversation will be recorded with your permission.

**Explain that there are 3 broad themes the conversation is going to cover:**

- If anything has changed for you / (patient's name) since our last conversation
- Your experiences of Just in Case drugs
- What you consider is important in supporting your / (patients name) comfort now

**Semi-structured interview questions**

***Prompt: turn recorders on***

1. **Can you start by telling me what has changed for you / (patient's name) since we last met?**

*Prompts:*

- *Have you / has (patient's name) had problems with any symptoms in the last few days?*
- *Which healthcare professionals are now involved in your / (patient's name) care?*

**Move on to themes going to cover: Thank you, that is really useful to understand. I would now like to ask about your opinions and experiences of "Just in Case drugs".**

2. **Since we last met have you discussed your Just in Case drugs with any healthcare professionals? If yes, would you mind telling me about this experience?**

**Go to question 7. If drugs have not been used.**

3. **If JIC drugs administered: Could you mind telling me about your experiences of having some of the drugs given by a nurse or doctor?**

*Prompts:*

- *How did the drugs come to be used?*
- *How did you find the process of getting someone to come and give the drugs?*
- *If they were called for help, how quick were professionals in responding / coming?*
- *Who made the decision to give the drugs?*
- *Did you feel you were involved in the decision to give the drugs?*

4. **If JIC drugs administered: Was any information about the drugs being administered discussed with you?**

*Prompts:*

- *Did a nurse or doctor explain what they were for and their effects before they administered them?*
- *Did anyone explain about the drugs and their effects after they were administered?*

- *Was the information and the way it was supplied helpful?*
- *Was there information that was not helpful?*
- *Was there information not talked about that you would have found helpful to know?*
- *If a syringe driver was set up, did anyone discuss what it was for with you?*

5. **If JIC drugs administered: Did the drugs help or hinder you / (patient's name) in any way?**

6. **If JIC drugs administered: How do you feel about these drugs being used?**

Prompts:

- *If a syringe driver was set up, how did you feel about this?*
- *Does the participant / do the participants expect Just in Case drugs to be used again?*
- *How do they feel about that?*

7. **If JIC drugs not administered: How do you feel about the Just in Case drugs being prescribed but not used?**

*Prompts:*

- *How do you feel about having the Just in Case drugs in the house for this length of time?*
- *Do you expect them to be used in the future?*
- *How do you feel about this?*

8. **Has having the Just in Case drugs prescribed affected how you see the future, or what is likely to happen in future?**

*Prompts:*

- *If yes, how has it affected how you see the future?*
- *Are you happy to have drugs in the house 'Just in Case' or does this cause you anxiety?*

**Thank you for sharing your thoughts and experiences.** I'd like to explore your / (patients name) care beyond just focusing on drugs to understand what's important to you.

9. **What is important in supporting your / *(patient's name)* comfort now?**

*Prompts:*

- *Follow the participant/s prompts, exploring their care wishes and understanding of their situation.*
- *If the participant/s raise that (patient's name) is becoming more unwell, give them the opportunity to talk about what they expect to happen, their hopes and concerns.*
- *Do you have any hopes or concerns about your / (patient's name) comfort in the future?*
- *Do you feel Just in Case drugs will be helpful in enabling you / (patient's name) to stay at home? (if the participant/s raise this as something that is important to them)*

10. **I think that's everything I wanted to talk about, do you have anything else you would like to say or pressing thoughts that I have not asked you?**

**Close the interview**

### Bereaved Informal Caregiver Participant Interview Schedule Three

**JUST IN CASE DRUGS STUDY**

**INTERVIEW SCHEDULE: BEREAVED INFORMAL CAREGIVER (FAMILY OR FRIEND) INTERVIEW THREE**

Note: 1. Questions will be continually reshaped in response to data from previous interviews.
2. This follow-up interview will build on and refer to what was established in the previous ones.

**Introduction:**

- Check the participant is happy to take part in the research conversation.
- The purpose of this follow-up research conversation is to ask about your views and experiences of 'Just in Case drugs' and their role in (patient's name) care.
- Clarify that there are no right or wrong answers.
- We can pause or stop the conversation at any time, please just say if you would like to.
- Please ask for explanations of any questions whenever you want.
- Our conversation will be recorded with your permission.

**Explain that there are 3 broad themes the conversation is going to cover:**

- If (patient's name) had any Just in Case drugs administered before they died
- Your experiences of the Just in Case drugs if they were used
- What was important to you in supporting *(patient's name)* in their last days of life

**Semi-structured interview questions**

*Prompt: turn recorders on*

1. **Would you be happy to talk me through your experiences of (patient's name) care in their last few weeks of life? (this will be tailored to the patient's situation)**

**Move on to themes going to cover: Thank you, it is incredibly helpful to understand your experiences. I would now like to ask about your opinions and experiences of the "Just in Case drugs".**

2. **Did (patient's name) have any of Just in Case drugs administered before they died?**

*Prompts:*

- *How did the drugs come to be used?*
- *How did you find the process of getting someone to come and give the drugs?*
- *If they were called for help, how quick were professionals in responding / coming?*
- *Who made the decision to give the drugs?*
- *Did you feel you were involved in the decision to give the drugs?*

**Go to question 6. if drugs have not been used.**

3. **If JIC drugs administered: Was any information about the drugs being administered discussed with you?**

*Prompts:*

- *Did a nurse or doctor explain what they were for and their effects before they administered them?*
- *Did anyone explain about the drugs and their effects after they were administered?*
- *Was the information and the way it was supplied helpful?*
- *Was there information that was not helpful?*
- *Was there information not talked about that you would have found helpful to know?*
- *If a syringe driver was set up, did anyone discuss what it was for with you?*

4. **If JIC drugs administered: Did the drugs help or hinder (patient's name) in any way?**

5. **If JIC drugs administered: How did you feel about these drugs being used?**

Prompts:

- *If a syringe driver was set up, how did you feel about this?*

6. **If JIC drugs not administered: How do you feel about the Just in Case drugs being prescribed but not used?**

*Prompts:*

- *How did you feel about having the Just in Case drugs in the house?*

7. **Did having the Just in Case drugs prescribed affect what you expected to happen to (patient's name) in their last days of life?**

*Prompts:*

- *Did you expect them to be used? - How did you feel about this?*
- *Were you happy to have drugs in the house 'Just in Case' or did this cause you any concern?*
- *Was (patient's name) happy to have the drugs in the home?*

8. **What was important to you in supporting *(patient's name)* in their last days of life?**

*Prompts:*

- *Did you have any hopes or concerns about (patient's name) comfort in their last days of life?*
- *Did you discuss your hopes or concerns with anyone?*
- *Overall, do you feel having the Just in Case drugs prescribed was helpful?*

9. **I think that's everything I wanted to talk about, do you have anything else you would like to say or pressing thoughts that I have not asked you?**

**Close the interview**

### Clinician Participant Interview Schedule

#### JUST IN CASE DRUGS STUDY

#### INTERVIEW SCHEDULE: CLINICIAN

Note: questions will be continually reshaped in response to data from previous interviews.

**Introduction:**

- Introduce yourself.
- Discuss the purpose of the study (the purpose of this research is to find out about patients and their family / friends opinions and experiences of 'Just in Case drugs', and if these change over time. We are also interested in understanding patients **and informal caregivers** involvement in the discussion and decision about JIC drugs. The focus of the interview will be on specific patient cases and it may help to have their medical records to hand to jog your memory)
- Clarify that there are no right or wrong answers. This research is to understand patients, informal caregivers, and healthcare professionals' perspectives and if these are similar or differ.
- We can pause or stop the interviews at any time, please just say if you would like to.
- Please ask for explanations of any questions whenever you want.
- Interviews will be recorded with your permission.
- All participant information collected during the research will be kept strictly confidential. The only situation in which we foresee that confidentiality might be broken would be if information is identified during the research that causes concern for the welfare of patients or others.

**Explain that there are 3 broad themes interview is going to cover:**

- How you see your role in looking after terminally ill people
- Your experiences of conversations with (patients name) and their family / friend around the prescribing and use of Just in Case drugs.

- The role of Just in Case drugs in the patient's care.

**Semi-structured interview questions**

*Prompt: turn recorders on*

1. **Can you start by telling me a little about how long you have been a (name of professional title)?**

2. **Can you tell me a bit about your role in looking after terminally ill people?**

**Move on to themes going to cover: Thank you, that is really useful to understand. I would now like to ask about one of the patient cases I am exploring as part of my research.**

3. **Could you tell me about your experiences of conversations with (patient name) and their informal caregiver (family / friend) around prescribing the Just in Case drugs?**

*Prompts:*

- *Can you recall the discussion with the patient and / or their informal caregiver about prescribing the drugs?*
- *If yes: how did it go? – were there any barriers or facilitators to conversations?*
- *Did they talk about the drugs or ask further questions about them once they were in the home?*
- *Do you think they clearly understood the purpose and significance of the Just in Case drugs?*
- *How do you think the patient / informal caregiver felt about having Just in Case drugs in the home?*

4. ***Did you take the initiative to prescribe the drugs or was it in response to another healthcare professional's intervention?***

*Prompts:*

- *Were there any events or changes that prompted the decision to prescribe drugs?*

**Go to question 9. if drugs have not been used.**

5. **(If drugs administered) Can you remember why drugs were administered?**

*Prompts:*

- *Which ones and for how long were they given?*
- *Who administered the drugs?*
- *Did any events or changes prompt the decision to give the drugs?*

6. ***(If drugs administered)* Can you recall if you or other professionals had any conversations about administering the Just in Case drugs with the patient or their informal carer?**

*Prompts:*

- *What was discussed?*
- *How did it go? – were there any barriers or facilitators to conversations?*
- *If no discussions, were there reasons why they did not happen?*

7. ***(If drugs administered)* Were the patient and their informal caregiver involved in the decision to administer the Just in Case drugs?**

*Prompts:*

- *If no, were there reasons why they were not involved in the decision to use the drugs?*

8. **Do you think the patient and their informal caregiver clearly understood the purpose of the drugs being administered?**

*Prompts:*

- *How do you think the patient / informal caregiver felt about having the drugs administered?*

9. ***(If drugs not administered)* Could you explain the reasons why drugs were not used?**
10. **I think that's everything I wanted to talk about, do you have anything else you would like to say or final thoughts that I have not asked you?**

**Close the interview**
