## Supplemental file 2 for "Mapping patient journeys: a novel method to explore patient and carer experiences of injectable anticipatory medication care in the community and identify opportunities for improvement"

***Supplementary file 2 – Narratives for additional patient cases***

***(Cases 1, 2 and 7 were used as the personas in the main paper. Further details in table 1 of main paper)***

**Patient case 3: Dave**

Dave lived at home with his wife, with his daughters living nearby. Dave was slowly deteriorating due to chronic kidney failure but continued to enjoy doing crosswords and being at home surrounded by his family. Following a six-week stay in hospital, Dave was reviewed at home by a GP, who explained to Dave that he was nearing the end of his life. Dave was prescribed anticipatory medication by his GP, and his daughter went to collect them. A district nurse then came around to check everything was in order and explained what the medications were for. This was very reassuring to the family, who trusted the nurses to do what was best. While Dave wasn't in pain at the time, the family found the presence of medication comforting, knowing that if he did experience pain, it could be treated.

Dave was an anxious person who appeared to prefer not knowing his prognosis and treatment and allowed his family to take care of that side of things. However, Dave was adamant that he did not want to be admitted to hospital. Dave's daughters were reassured that the end-of-life medication could help to prevent this from happening. Dave's daughter was concerned about whether the drugs, if not used, would go to waste or if there was a way to recycle them. At the final interview Dave was still at home and the anticipatory medications had not been used.

**Case 4: Helen**

Helen lived in a care home following a stroke. At the beginning of the year, she began to deteriorate, eventually becoming unable to mobilise. Her daughter Alice pushed for a blood test, which ultimately led to a diagnosis of terminal cancer. Helen declined any treatment that would require her to go to hospital. However, she was never consulted about the "just in case" medications that were prescribed. Alice couldn't recall any conversation with the doctor about the medications being prescribed or any explanation provided about them. When Alice was told by the care home's team leader that the medication had arrived, Alice didn't question it as it made sense to her. Alice had previous experience with "just in case" medication, having advocated for their use with her father.

When Helen became distressed, Alice felt it was necessary to start giving the anticipatory medication, and felt this was supported by the care home staff. However, the first district nurse who visited didn't think this was necessary. Alice was accustomed to advocating for her mother and pushed for the medication to be used. A second district nurse arrived, who agreed with Alice. Helen received two injections of the sedative, and by the third day, a syringe driver was set up, and from then onwards remained peaceful. Alice was relieved that she was able to be with her mother in her final week. Looking back, Alice felt certain that her active role in advocating for Helen's comfort had made all the difference in her mother's final days.

**Case 5: Hugo**

Hugo had been seen by the district nurses regularly for two years but had otherwise been managing at home with the help of his daughter, Emily, who lived next door. Emily arranged a GP appointment after his heart failure deteriorated and he became increasingly short of breath. Following the video GP appointment, the district nurse asked Emily if it would be okay to request anticipatory medication and refer him to palliative care. Because she had already had these discussions with the GP, she wasn't distressed by the conversation and agreed. She was adamant she didn't want him to go to hospital, however she felt that if she had said no to the medication or palliative care referral, her wishes would have been respected.

Emily and her husband could help during the day as they were both working from home. Emily expressed that, as she is a nurse, the drugs and their uses were not explained to her in detail, although she hoped that if she had not known about the drugs, they would have been explained to her. Emily's understanding and perception of the drugs were influenced by her experience of having to advocate for their use with her mother at the end of her life previously, and she did not want her father to experience the same distress at the end of his life.

Emily administered a lorazepam tablet (supplied in the anticipatory medication kit) when Hugo became agitated, and although she found it useful for reducing his agitation, she did not like how sedated it made him. She limited its use but did find it reassuring having the injectable medication in the house. She hoped that Hugo would die in his sleep without needing to use them again. At the final interview Hugo had moved into a nursing home and the drugs had not been used again.

**Case 6: Louise**

Louise had cancer. She received radiotherapy, but after developing sepsis, was told she had weeks left to live. She suffered from bloating and stomach pains and tried to manage this with herbal medicine, alongside medication for liver problems. She found the fatigue frustrating, as she was previously very active and enjoyed cycling. However, she was able to take the dog for a walk every day to keep exercising. She found the support from dietitians, the palliative care team, and the district nurses very helpful. Louise actively researched ways to help manage her symptoms. She also took morphine to help with the pain but didn't want to increase the dose if it made her drowsy, as she wanted to remain active. Louise was supported by her partner and son.

Louise was prescribed anticipatory medication after being discharged from hospital and assumed it was the GP who prescribed them but was not sure. She was informed by phone that the medications would be delivered by the pharmacy. However, no one explained what the drugs were specifically for; Louise assumed they were for helping to control end of life symptoms as she didn't want to go into hospital. Louise had previously gone to the emergency department for pain relief, so was relieved that having the anticipatory medications meant that she could obtain pain relief at home. Louise used the "just in case" medication on a few occasions, for stomach and back pain, which the morphine helped alleviate. Unexpectedly Louise was told she could have an operation to remove her cancer, which changed the prognosis of her disease. Her view of the drugs changed from seeing them as end-of-life medication to pain and symptom control medication whilst she awaited surgery.

**Case 8: Ruth**

Ruth lived with her grandson Mark, supported by district nurses and hospice at home nursing team. She had been given a prognosis of a few months following a cancer relapse and was deteriorating but was pleased to be able to spend time with her grandson. Ruth was prescribed anticipatory medication by a GP following a hospital admission for a medical procedure. Mark advocated for this to be done quickly so that Ruth's discharge from the hospital wasn't delayed. The doctor didn't explain what the medication was for, but the district nurses and palliative care team explained to Mark that it was to keep Ruth comfortable. The district nurses also explained this to Ruth, but Mark reiterated everything afterwards to make sure Ruth understood fully. Mark went to collect the paperwork from the surgery, and then the medication was delivered to them.

The anticipatory medication was used for pain relief when Ruth's pain was not controlled with her usual medication. Their experience of getting the district nurses involved and the anticipatory medication administered was positive. However, it got to the point where they were having to call out the district nurses every day to give the injectable medication. The district nurses always arrived within an hour, and Mark found them very helpful and responsive. Mark also found that the medication always worked within 30 minutes. Ruth had a syringe driver for 24 hours but decided to have it removed, as she didn't like having another thing attached to her, even though it was more effective at controlling her symptoms. Eventually, the decision was made for Ruth to be admitted to her local hospice. Mark found the most frustrating part was reordering medication. He would have to get it re-prescribed by a GP, then go to collect it, only to find it wasn't available. This led to a lot of back-and-forth, which was frustrating, particularly as they live rurally. Mark found it more effective when the district nurses called the surgery to emphasise the urgency of the new prescriptions.

**Case 9: Joe**

Joe lived with Kim, his wife, who helps care for him but was limited in what she could do as she also has a life limiting condition. Joe enjoyed working on and driving his three cars and just two weeks prior to his research interview, he was still driving. However, his condition deteriorated rapidly, and he was in a lot of pain. This limited his ability to walk. He also suffered from morphine-related constipation, which caused additional pain and was not well-controlled with laxatives. Joe viewed the anticipatory medication as a safety net in case he needed to call the district nurse out during the night, when it might be harder to get hold of the medication to control his pain and prevent him from having to go to the hospital. He picked up the anticipatory medication from his local pharmacy, which he found easy. The doctor who prescribed the medication didn't explain what they were for, and although he had a discussion with the district nurse, she also didn't provide details about what the drugs were for. Joe planned to ask for more information the next time he saw his palliative care nurse. Joe and his wife felt that it was their choice to have the medication, which also helped Kim, Joe's partner, to relax, as it meant she was not worried about needing to drive Joe to the emergency department. At the final interview Joe was still at home and the anticipatory medications had not been used.

**Case 10: Abby**

Abby lived at home with her husband, with support from district nurses, and was about to have carers coming in to help. She had been slowly deteriorating due to a respiratory disease which limited her mobility, and required home oxygen therapy, both at rest and when active. Even talking required high levels of supplemental oxygen. While this was necessary for managing her breathlessness, it meant she could no longer cook, which was a hobby she enjoyed before. Abby went to a local hospice for day therapy, and one of the hospice nurses suggested that her GP prescribe anticipatory medication, which Abby agreed to. The nurse then contacted her GP, and anticipatory medication was prescribed. Abby's husband picked it up from the pharmacy. The GP then visited and explained what all the medication was for, which Abby found extremely helpful.

Abby felt that the GP, hospital and hospice staff all worked well together. However, it helped that Abby was able to advocate for herself and took paper notes and blood test results to her different doctor appointments. She understood that the medication was intended to make her last hours more comfortable, but she also expressed that it was not meant to over-sedate her. While she knew her prognosis was very uncertain, she felt the anticipatory medication would help make the final stages of her life easier for her family, as she hoped she would be more peaceful. This was comforting for her, as she was concerned about the effect of her disease on her family. At the final interview Abby was still at home and the anticipatory medication had not been used.

**Case 11: Dylan**

Dylan lived at home with his wife Freya in a very rural location. He had been unwell with cancer for two years but had deteriorated over the past three months, experiencing poorly controlled pain and nausea. Dylan was grateful that he had Freya to advocate on his behalf; otherwise, he didn't think he would be able to navigate the system. Dylan was going regularly to the palliative care day unit, where a palliative care nurse recommended anticipatory medications. Both Dylan and Freya agreed that this would be helpful. They had a discussion with the palliative care nurse about the medication, but not with the GP who prescribed it. They found it took a long time for the GP surgery to act on the advice of the palliative care nurse.

A district nurse suggested starting Dylan with a syringe driver. However, they had difficulty getting the medication restocked after this was started, and on one occasion they were left without any before the weekend. They had similar problems obtaining stocks of water for injection and further needles, which were not delivered with the original medication. During Dylan's final few weeks of life, there were ongoing issues with the anticipatory medication prescriptions. On the day Dylan died, there was a significant delay in giving him medication, as the district nurse had to go back to the surgery to get the prescription chart adjusted. Freya thought that many of the delays were due to miscommunication between the GP surgery and the district nurses administering his care. However, when the medication was eventually administered, it appeared to help with Dylan's pain. After Dylan's death, the leftover anticipatory drugs were collected by someone from the nursing team, but most of Dylan's other prescription medication remained unopened in the house. Freya was frustrated that they couldn't be recycled. She also had nagging doubts about how effective the medication was in relieving Dylan's pain in his last hours of life.
