## Supplemental file 3 for "Mapping patient journeys: a novel method to explore patient and carer experiences of injectable anticipatory medication care in the community and identify opportunities for improvement"

### Supplementary file 3 - Initial mapping process for Liam

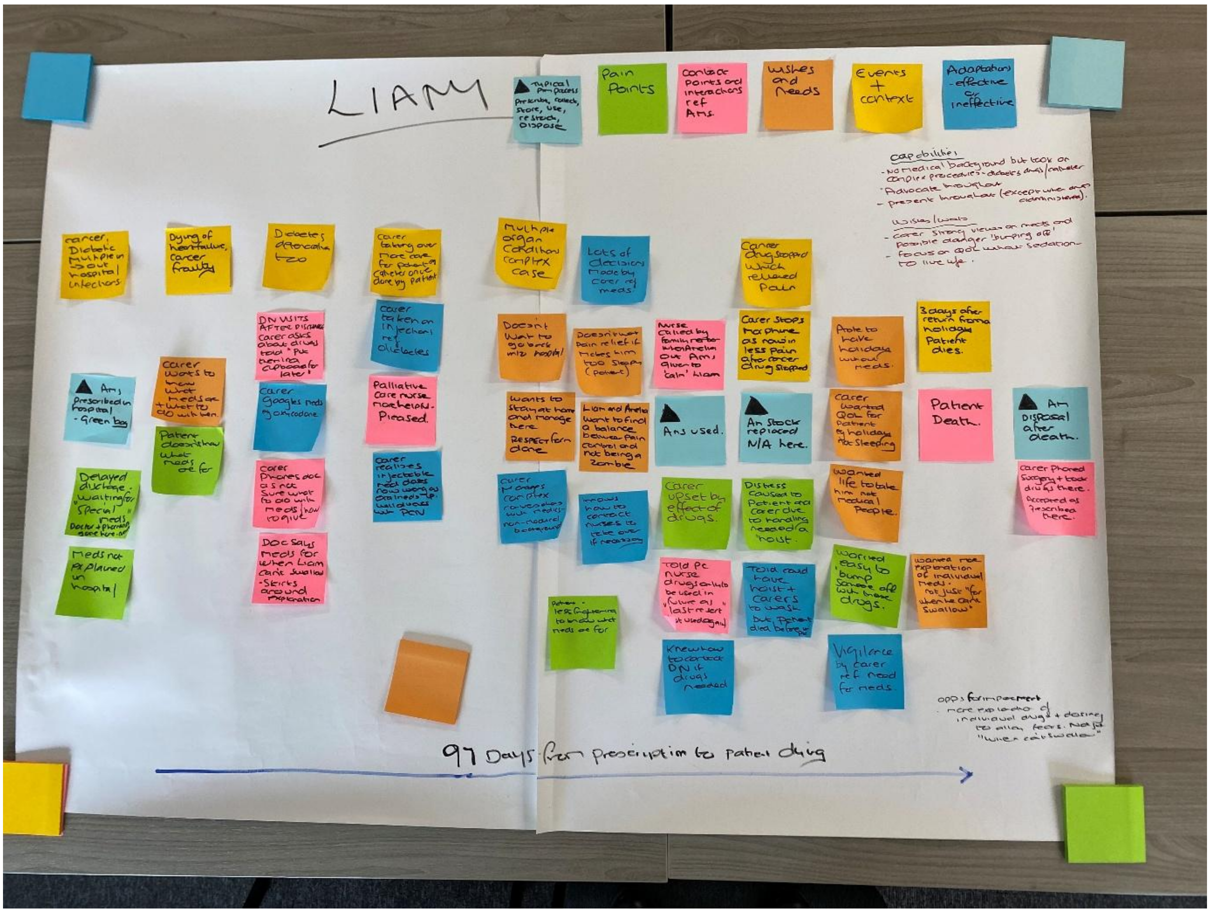


*Fig 1: Initial journey mapping process for Liam using sticky notes*

Initial visualisations using sticky notes were completed for each persona and were subsequently condensed and professionally illustrated for inclusion in the paper.

Each journey comprised the intended pathway with anticipatory medications (prescribe, dispense, store, use (or not), dispose) – shown as pale blue sticky notes with triangles. The remaining colours represent patient and carer experiences related to the intended pathway, including patient contexts and characteristics. Experiences related to components of the pathway were mapped as follows.

- Pale Blue with triangles – intended pathway with anticipatory medications (AMs)
- Pink – patient experienced pathway with anticipatory medications including interactions
- Orange – patient or informal carer wishes and/or needs
- Green – perceived pain points
- Yellow – events and context related by participants
- Darker Blue – patient or carer adaptations - both effective and ineffective
- Capabilities were also listed
